# Clinical and genetic analysis of Costa Rican patients with Parkinson’s disease

**DOI:** 10.1101/2020.09.29.20202432

**Authors:** Gabriel Torrealba-Acosta, Eric Yu, Tanya Lobo-Prada, Javier Ruiz-Martínez, Ana Gorostidi-Pagola, Ziv Gan-Or, Kenneth Carazo-Céspedes, Jaime Fornaguera-Trías

## Abstract

**Background:** Parkinson’s disease (PD) involves environmental risk and protective factors as well as genetic variance. Most of the research in genomics has been done in subjects of European ancestry leading to sampling bias and leaving Latin American populations underrepresented.

**Objective:** We sought to phenotype and genotype Costa Rican PD cases and controls.

**Methods:** We enrolled 118 PD patients with 97 unrelated controls. Collected information included demographics, exposure to risk and protective factors, motor and cognitive assessments. We sequenced coding and untranslated regions in familial PD and atypical parkinsonism-associated genes including *GBA, SNCA, VPS35, LRRK2, GCH1, PRKN, PINK1, DJ-1, VPS13C, ATP13A2*.

**Results:** Mean age of PD probands was 62.12 ± 13.51 years, 57.6% were male. Prevalence of risk and protective factors reached 30%. Physical activity significantly correlated with better motor performance despite years of disease. Increased years of education were significantly associated with better cognitive function, whereas hallucinations, falls, mood disorders and coffee consumption correlated with worse cognitive performance. We did not identify an association between tested genes and PD or any damaging homozygous or compound heterozygous variants. Rare variants in *LRRK2* were nominally associated with PD, six were located between amino acids p.1620-1623 in the C-terminal-of-ROC (COR) domain of LRRK2. Nonsynonymous *GBA* variants (p.T369M, p.N370S, p.L444P) were identified in three healthy individuals. One PD patient carried a pathogenic *GCH1* variant, p.K224R.

**Conclusion:** This is the first study that reports on sociodemographic, risk factors, clinical presentation and genetics of Costa Rican patients with PD.

## 1. Introduction

Parkinson’s disease (PD) is a complex and heterogeneous movement disorder caused by a progressive degeneration of dopaminergic neurons. Main clinical motor symptoms associated with PD include tremor, rigidity, bradykinesia and postural imbalance [1]. Years before motor symptoms are manifested there can be prodromal non-motor key features that include REM sleep disorders, anosmia and constipation [2]. Its pathophysiology involves environmental factors as well as genetic variance, which provide insight into its molecular pathogenesis. Among environmental factors that contribute to PD risk include pesticide and herbicide exposure, welding and well water consumption. There are also protective factors such as smoking, coffee consumption and performing physical activity that may reduce the risk of developing PD [3].

Since the description of PD associated mutations in the *SNCA* [4], other genes have been linked to autosomal dominant (AD) forms of familial PD, including *LRRK2* and *VPS35*. In addition, there are clinically and genetically diverse early-onset (EO) autosomal-recessive (AR) forms of PD with associated genes like *PRKN, PINK1, and DJ-1* that exhibit phenotypes similar to idiopathic PD; while other associated genes such as *VPS13C, ATP13A2* combine atypical features of parkinsonism like dystonia and early cognitive impairment, along with a poor response to levodopa [5]. Large-scale genome-wide association studies (GWASs) have established different *GBA* locus as strong risk factors for PD both in homozygous and heterozygous state; displaying a phenotype similar to idiopathic PD, yet with a faster rate of progression of cognitive and motor decline [6].

Most of the work and research in genomics has been done in subjects of European ancestry leading to sampling bias and leaving large populations underrepresented [7]. Few genetic studies have been conducted shedding some light on the genotype and phenotype of Latin American populations [8-20]. Current efforts to include Latin Americans involve the LARGE-PD initiative of extending large scale genotyping to these cohorts [21]. In this study, we sought to clinically characterize PD patients of Costa Rican origin and to sequence familial PD and atypical parkinsonism-associated genes in Costa Rican PD cases and controls.

## 2. Materials and Methods

### 2.1. Study subjects

We enrolled 118 consecutive unrelated PD patients (68 males, 50 females) with 97 unrelated controls (28 males, 69 females), matched according to age and gender whenever possible. Thirty-five patients (16.28%) reported having a relative (≤ 2° degree) with any sort of movement disorder; of those, 21 (9.77%) had a formal PD diagnosis. All of the subjects resided and were originated from Costa Rica and were recruited at the Movement Disorders Unit of the Department of Neurology, Hospital San Juan de Dios, Caja Costarricense de Seguro Social. All patients fulfilled Gelb criteria for the clinical diagnosis of PD; while controls had no signs or personal history of any neurodegenerative disease and were mainly the spouses of the PD cases. We gathered information concerning work and educational status as well as history of exposure to risk and protective factors of PD. We further obtained detailed information on PD history, comorbidities and antiparkinsonian treatment. Additionally, motor disability of the patients was evaluated by means of the Unified Parkinson’s Disease Rating Scale (UPDRS), Hoehn & Yahr (H&Y) and Schwab & England (S&E) scales. Cognitive status was assessed using the Montreal Cognitive Assessment (MoCA) test.

### 2.2. Genetic analysis

#### Genetic analysis

Molecular inversion probes were used to sequence coding and untranslated regions in familial PD and atypical parkinsonism-associated genes including *GBA, SNCA, VPS35, LRRK2, GCH1, PRKN, PINK1, DJ-1, VPS13C, ATP13A2* at McGill University with Illumina HiSeq 4000 as previously described [22]. The full protocol can be found at https://github.com/gan-orlab/MIP_protocol. All sequences were aligned using Burrows-Wheeler Aligner (BWA) using the reference genome hg19 [23]. Genome Alignment Took Kit (GATK v3.8) was used to call variants and perform quality control and ANNOVAR was used to annotate each variant [24, 25]. Exons 10 and 11 of *GBA* were sequenced using Sanger sequencing as previously described [26], and *GBA* variants in other exons were also validated using Sanger sequencing.

#### Quality Control

All samples and variants were filtered based on standard quality control process as previously reported [27]. In brief, variants were separated into common and rare by minor allele frequency (MAF) in the cohort. Rare variants (MAF < 0.01) with a minimum depth of coverage of >30x were included in the analysis, along with common variants (MAF >= 0.01) with >15x coverage.

Variant calls with a genotype frequency of less than 25% of the reads or genotype quality of less than 30 were excluded. Samples and variants with more than 10% missingness were also excluded.

### 2.3. Statistics

We used Stata® (version 14) for the statistical analysis of sociodemographic and clinical variables. Normally distributed variables are reported as mean with its standard deviation (SD), whereas continuous but non-normally distributed variables are reported as median with the 25^th^ and 75^th^ percentile values (Q1-Q3). Normally distributed variables were compared with paired or unpaired t-tests, while non-normally distributed variables were compared with Mann-Whitney U test or Wilcoxon match-paired signed-rank test. Frequencies were compared with χ^2^ and Fisher’s exact test. Tests were 2-tailed, and significance was set at *p*<0.05. We modeled through linear regression the association between demographic and clinical variables with the severity of the disease, as indexed by UPDRS and MoCA.

For genetic analysis, common and rare variants were analyzed separately. Association of common variants was tested using logistic regression adjusted for age and sex in PLINK v1.9. For rare variants’ analysis, we examined the burden of rare variants in each gene using optimized sequence Kernel association test (SKAT-O) adjusted for age and sex [28]. Rare variants were separated into different categories based on their potential pathogenicity to examine specific enrichment in different variant subgroup as described previously [29]: 1) variants with Combined Annotation Dependent Depletion (CADD) score of ≥12.37 (representing the top 2% of potentially deleterious variants) [30]; 2) regulatory variants predicted by ENCODE [31]; 3) potentially functional variants including all nonsynonymous variants, stop gain/loss variants, frameshift variants and intronic splicing variants located within two base pairs of exon-intron junctions; 4) loss-of-function variants which includes stop gain/loss, frameshift and splicing variants; 5) only nonsynonymous variants. Bonferroni correction for multiple comparisons was applied as necessary.

This study was approved by the Ethics Committee of Hospital San Juan de Dios, Caja Costarricense de Seguro Social (CLOBI-HSJD #014-2015) and the University of Costa Rica (837-B5-304). Written informed consent was obtained from all participants.

## 3. Results

### 3.1. Sociodemographic and clinical variables

At enrollment, PD probands had a mean age of 62.12 ± 13.51 years (range 25-86), and the mean age at onset was 54.62 ± 13.54 (range 16-83) years. Male PD patients comprised 57.63% of the sample with a mean age of 62.75 ± 12.17, while females reported a mean age of 61.26 ± 15.22 years at recruitment. Despite a significantly larger proportion of the male PD patients reported current or previous jobs involving agricultural activities (19.40% male, 2.08% female, *p*=0.01), the mean number of years of education of these men was significantly higher than women (10.74 ± 3.81 vs 8.86 ± 4.01, *p*=0.03). Table 1 details subjects’ baseline characteristics along with the prevalence of exposure to main risk and protective factors for PD. Most of the risk and protective factors were more prevalent in men. Tables 1 and 2 detail the frequency of clinical manifestations as well as the standardized scale scores reported for PD cases. The most frequent initial symptomatology of the patients included: resting tremor (71.30%), rigidity (24.07%) and pain (10.19%). Most of the patients had asymmetric onset (94.12%) and a good response to levodopa (89.11%). Other frequently reported motor features comprised dystonia (46.08%), falls (39.22%), and dysphagia (36.27%). Common non-motor manifestations such as hiposmia, sleep disorders and depressive/anxious mood were seen in more than 50% of the PD cases. Overall median score of UPDRS ‘ON’ was 36 (22-60), most of our patients were graded in the ‘2.5’ and ‘3’ categories of the H&Y scale with a median for S&E score of 80% (80-90%). The median value for the MoCA test was 22 (17-25). There were no statistically significant differences between sex, regarding these scores.

**Table 1.** Baseline characteristics with prevalence of risk and protective factors for PD in study subjects, with sex comparison.

|  | **Men**<br>**96 (44.65%)** | **Women**<br>**119 (55.35%)** | **Total**<br>**(n=215)** | **P** |
| --- | --- | --- | --- | --- |
| **Condition** |  |  |  | **<0.001** |
| **Cases** | 68/118 (57.63%) | 50/118 (42.37%) | 118/215 (54.88%) |  |
| **Controls** | 28/97 (28.87%) | 69/97 (71.13%) | 97/215 (45.12%) |  |
| **Age of onset of PD (mean ± SD)** | 54.74 ± 12.03 | 54.46 ± 15.49 | 54.62 ± 13.54 | 0.91 |
| **Age of recruitment (mean ± SD)** | 62.75 ± 12.17 | 61.26 ± 15.22 | 62.12 ± 13.51 | 0.10 |
| **Years of education<sup>¶</sup> (mean ± SD)** | 10.74 ± 3.81 | 8.86 ± 4.01 | 9.94 ± 3.98 | **0.03** |
| **Agricultural activities<sup>¶</sup>, n (%)** | 13/67 (19.40%) | 1/48 (2.08%) | 14/115 (12.17%) | **0.01** |
| **Risk factors<sup>¶</sup>, n (%)** |  |  |  |  |
| **Pesticides** | 24/67 (35.82%) | 10/48 (20.83%) | 34/115 (29.57%) | 0.10 |
| **Herbicides** | 26/67 (38.81%) | 9/48 (18.75%) | 35/115 (30.43%) | **0.03** |
| **Welding** | 22/65 (33.85%) | 3/48 (6.25%) | 25/113 (22.12%) | **<0.001** |
| **Heavy metals** | 11/64 (17.19%) | 2/48 (4.17%) | 13/112 (11.61%) | **0.04** |
| **Non-potable water** | 29/66 (43.94%) | 19/48 (39.58%) | 48/114 (42.11%) | 0.70 |
| **Cardiovascular** | 41/58 (70.69%) | 32/43 (74.42%) | 73/101 (72.28%) | 0.82 |
| **Years of exposure, median (Q1-Q3)** | 8 (1-20) | 5 (1-19) | 8 (1-20) | 0.36 |
| **Protective factors<sup>¶</sup>, n (%)** |  |  |  |  |
| **Smoking** | 36/67 (53.73%) | 9/48 (18.75%) | 45/115 (39.13%) | **<0.001** |
| **Coffee** | 61/65 (93.85%) | 44/48 (91.67%) | 105/113 (92.92%) | 0.72 |
| **Alcohol** | 43/67 (64.18%) | 9/48 (18.75%) | 52/115 (45.22%) | **<0.001** |
| **Physical activity** | 47/67 (70.15%) | 17/48 (35.42%) | 64/115 (55.65%) | **<0.001** |
| **UPDRS ‘on’ median (Q1-Q3)** |  |  |  |  |
| **I** | 2 (0.5-4) | 2 (0-5) | 2 (0-5) | 0.98 |
| **II** | 9 (3-17) | 8 (3-14) | 9 (3-16) | 0.57 |
| **III** | 23 (10-35) | 26 (14-37) | 23 (12-36) | 0.57 |
| **Total** | 35 (21-59) | 36.5 (23-60) | 36 (22-60) | 0.97 |
| **Hoehn & Yahr** |  |  |  |  |
| **1** | 5/59 (8.47%) | 9/43 (20.93%) | 14/102 (13.73%) | 0.36 |
| **1.5** | 7/59 (11.86%) | 6/43 (13.95%) | 13/102 (12.75%) |  |
| **2** | 12/59 (20.34%) | 5/43 (11.63%) | 17/102 (16.67%) |  |
| **2.5** | 14/59 (23.73%) | 9/43 (20.93%) | 23/102 (22.55%) |  |
| **3** | 17/59 (28.81%) | 8/43 (18.60%) | 25/102 (24.51%) |  |
| **4** | 3/59 (5.08%) | 4/43 (9.30%) | 7/102 (6.86%) |  |
| **5** | 1/59 (1.69%) | 2/43 (4.65%) | 3/102 (2.94%) |  |
| **Schwab & England %, median (Q1-Q3)** | 80 (80-90) | 90 (80-90) | 80 (80-90) | 0.33 |
| **MoCA test, median (Q1-Q3)** | 22.5 (18-25.5) | 22 (14-25) | 22 (17-25) | 0.38 |

<sup>a</sup>Information available only for patients and does not include controls.
Abbreviations: MoCA: Montreal Cognitive Assessment, PD: Parkinson's disease, Q1-Q3: first and third quartile, SD: standard deviation, UPDRS: Unified Parkinson's Disease Rating Scale.

**Table 2.** Clinical manifestations of PD cases, with sex comparison.

|  | **Men**<br>**68 (57.63%)** | **Women**<br>**50 (42.37%)** | **Total**<br>**(n=118)** | **P** |
| --- | --- | --- | --- | --- |
| **Initial symptoms, n (%)** |  |  |  |  |
| Resting tremor | 43/63 (68.25%) | 34/45 (75.56%) | 77/108 (71.30%) | 0.52 |
| Rigidity | 17/63 (26.98%) | 9/45 (20.00%) | 26/108 (24.07%) | 0.50 |
| Postural instability | 3/63 (4.76%) | 1/45 (2.22%) | 4/108 (3.70%) | 0.64 |
| Bradykinesia | 3/63 (4.76%) | 1/45 (2.22%) | 4/108 (3.70%) | 0.64 |
| Pain | 3/63 (4.76%) | 8/45 (17.78%) | 11/108 (10.19%) | **0.03** |
| **Symptoms during illness course, n (%)** |  |  |  |  |
| Resting tremor | 51/59 (86.44%) | 40/43 (93.02%) | 91/102 (89.22%) | 0.35 |
| Bradykinesia | 55/59 (93.22%) | 39/43 (90.70%) | 94/102 (92.16%) | 0.72 |
| Rigidity | 50/59 (84.75%) | 36/43 (83.72%) | 86/102 (84.31%) | 0.89 |
| Asymmetry | 57/59 (96.61%) | 39/43 (90.70%) | 96/102 (94.12%) | 0.24 |
| Levodopa response | 55/59 (93.22%) | 35/42 (83.33%) | 90/101 (89.11%) | 0.19 |
| Hallucinations | 14/59 (23.73%) | 8/43 (18.60%) | 22/102 (21.57%) | 0.63 |
| Orthostatism | 9/59 (15.25%) | 12/43 (27.91%) | 21/102 (20.59%) | 0.14 |
| Falls | 23/59 (38.98%) | 17/43 (39.53%) | 40/102 (39.22%) | 0.96 |
| Syncope | 1/59 (1.69%) | 2/43 (4.65%) | 3/102 (2.94%) | 0.57 |
| Dystonia | 28/59 (47.46%) | 19/43 (44.19%) | 47/102 (46.08%) | 0.84 |
| Dysphagia | 20/59 (33.90%) | 17/43 (39.53%) | 37/102 (36.27%) | 0.68 |
| Hyposmia | 34/63 (53.97%) | 23/47 (48.94%) | 57/110 (51.82%) | 0.70 |
| Constipation | 23/52 (44.23%) | 13/41 (31.71%) | 36/93 (38.71%) | 0.29 |
| Urinary symptoms | 7/52 (13.46%) | 5/41 (12.20%) | 12/93 (12.90%) | 0.86 |
| Sleep disorders | 76/95 (80.0%) | 49/68 (72.1%) | 125/163 (76.7%) | 0.26 |
| Insomnia | 22/52 (42.31%) | 16/37 (43.24%) | 38/89 (42.70%) | 0.93 |
| Vivid dreams | 26/52 (50.00%) | 13/37 (35.14%) | 39/89 (43.82%) | 0.20 |
| Psychiatric symptoms (depression or anxiety) | 40/63 (63.49%) | 29/47 (61.70%) | 69/110 (62.73%) | 0.85 |
| **Disease duration (years), median (Q1-Q3)** | 5 (3-10) | 5 (3-7) | 5 (3-9) | 0.18 |

Abbreviations: Q1-Q3: first and third quartile.

We were able to establish through multivariate linear regression modeling that an increased disease duration along with the presence of orthostasis, dysphagia and mood disorders significantly correlated with increased scores in total ON UPDRS. Furthermore, we found an interaction between performing regular physical activity and duration of disease, where despite having increased years of evolution, patients that performed regular physical activity still scored less in the total ON UPDRS (see Supplementary Figure 1). Additionally, lower scores in MoCA testing significantly correlated with increased age, coffee consumption and the presence of hallucinations, falls and mood disorders, whereas increased years of education correlated with better MoCA scores (see Supplementary Figure 2).

### 3.2. Quality of coverage and identified variants

The average coverage of the 10 genes analyzed in this study was >588X for all genes. The coverage per gene, and the percentage of nucleotides covered at >15X and >30X for each gene are detailed in Supplementary Table 1. There were no differences in the coverage across the samples (patients and controls). Overall, after quality control, we identified 163 rare variants (Supplementary Table 2 and 158 common variants (Supplementary Table 3 across all genes and all samples that were included in the analysis. Specific protein and DNA changes are listed in Supplementary Tables 4 and 5 for rare and common exonic variants, respectively.

### 3.3. Rare and common variants in Parkinson’s disease and parkinsonism-related genes

Burden and SKAT-O analyses did not identify an association of any of the tested genes and PD (Table 3) after correction for multiple comparisons, as expected given the small sample size. We also did not identify any PD patients with potentially damaging homozygous or compound heterozygous variants in any of these genes. Rare variants in *LRRK2* were nominally associated with PD, and 11 (9.2%) of patients carried a rare nonsynonymous variant, compared to four (4.1%) among controls. Interestingly, six of these rare nonsynonymous variants, all located between amino acids p.1620-1623 in the C-terminal-of-ROC (COR) domain of LRRK2, were found in six patients and none in controls (Table 4).

**Table 3.**
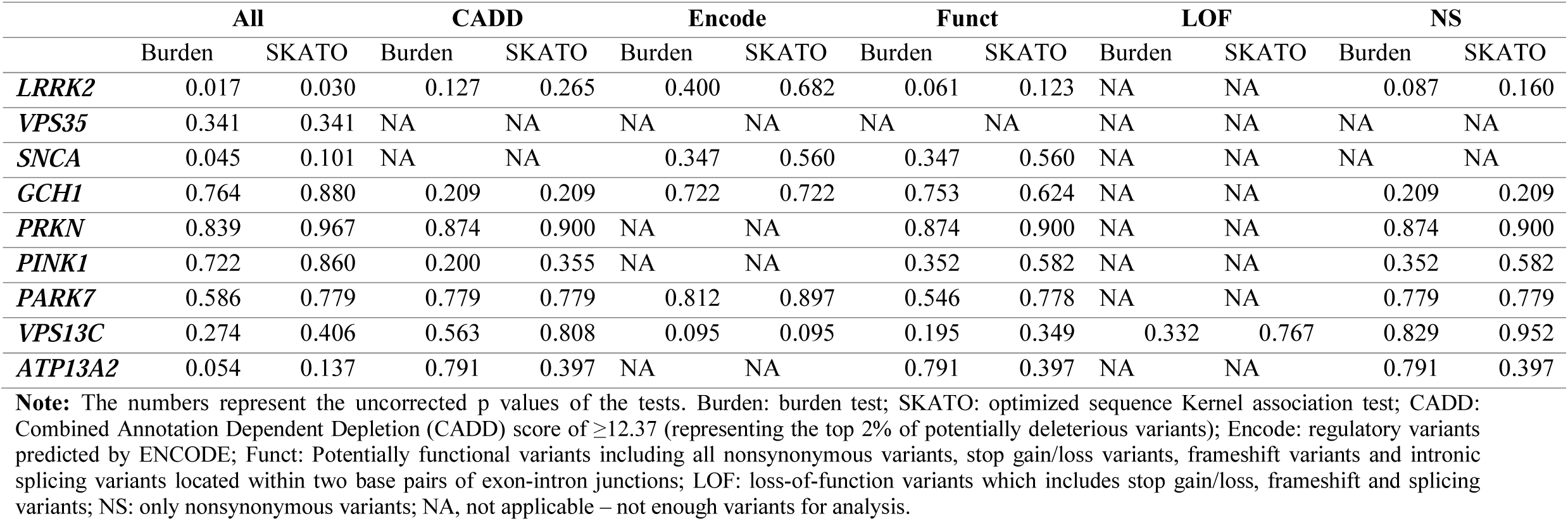
Burden and SKAT-O analyses with no significant association found of any of the tested genes and PD, after Bonferroni correction for multiple comparisons.

**Table 4.** Rare variants in *LRRK2* present in patients and controls. Six of these rare nonsynonymous variants (*), all located between amino acids p.1620-1623 in the COR domain of LRRK2, were found only in patients and not in controls.

| SNP location/rs number and nucleotide change | Detailed annotation of the variant (DA) | Status | Family History |
| --- | --- | --- | --- |
| **12:40709172:T:C** | intronic:LRRK2:NM_198578 | Control |  |
| **12:40709180:T:C** | intronic:LRRK2:NM_198578 | Affected |  |
| **12:40709181:T:C** | intronic:LRRK2:NM_198578 | Affected |  |
| **rs760912433:C:T** | exonic:nonsynonymous_SNV:LRRK2:NM_198578:exon34:c.C4856T;p.P1619L | Control |  |
| **12:40713821:A:G** | exonic:nonsynonymous_SNV:LRRK2:NM_198578:exon34:c.A4859G;p.K1620R | Affected |  |
| **12:40713824:A:G** | exonic:nonsynonymous_SNV:LRRK2:NM_198578:exon34:c.A4862G;p.H1621R | Affected | Mother:<br>• Epilepsy (type unknown) diagnosed in early adulthood.<br>• Dementia associated to rigidity, diagnosed at 67 years old. Death at 69 years old<br>• Retinitis Pigmentosa |
| **rs765275134:C:A** | exonic:nonsynonymous_SNV:LRRK2:NM_198578:exon34:c.C4863A;p.H1621Q | Affected |  |
| **12:40713826:C:A** | exonic:nonsynonymous_SNV:LRRK2:NM_198578:exon34:c.C4864A;p.P1622T | Affected | Father:<br>• Dementia (Type unknown), not associated to hallucinations or motor symptoms. Diagnosed at 74 yo, death at 84 yo. |
| **rs751492506:C:T** | exonic:nonsynonymous_SNV:LRRK2:NM_198578:exon34:c.C4865T;p.P1622L | Affected | Two Sisters (from both parents):<br>• Parkinson Disease diagnosed at ages 30 and 20 years old.<br>Maternal aunt:<br>• Bilateral hand tremor (described as intention tremor; does not have a definitive diagnosis).<br>Maternal Great-grandfather:<br>• Bilateral hand tremor (type unknown). |
| **12:40713828:T:C** | exonic:synonymous_SNV:LRRK2:NM_198578:exon34:c.T4866C;p.P1622P | Control |  |
| **12:40713829:A:G** | exonic;nonsynonymous_SNV:LRRK2:NM_198578:exon34:c.A4867G:p.K1623E | Affected | Mother:<br>• Cirrhosis (not associated to neurologic symptoms).<br>Maternal uncle:<br>• Parkinson Disease (diagnosed at 50 years old).<br>Maternal grandfather:<br>• Tremor in both hands (type unknown). |
| **rs73097447:A:C** | intronic:LRRK2:NM_198578 | Affected |  |
| **12:40716092:G:T** | intronic:LRRK2:NM_198578 | Affected<br>& Control |  |

Nonsynonymous *GBA* variants were identified in three individuals: p.T369M was identified in a male patient with age at onset of 48 years, p.N370S was identified in a healthy female individual recruited at the age of 78 years, and p.L444P was identified in a healthy female individual recruited at the age of 64. While we cannot rule out that these healthy individuals will develop PD in the future, it is unlikely that *GBA* variants have a major role in PD among Costa Rican patients. One PD patient carried a pathogenic *GCH1* variant, p.K224R, further emphasizing the role of this gene in PD.

In the analysis of common variants, none of the variants was associated with PD after correction for multiple comparisons (Supplementary Table 3), which set the corrected p value for statistical significance at p<0.00031. One nonsynonymous variant in *LRRK2*, p.I723V, was found with allele frequency of 0.01 in patients and 0.09 in controls (OR=0.11, 95% CI=0.02-0.52, *p*=0.005), yet this difference was not statistically significant after correction for multiple comparisons.

## 4. Discussion

### 4.1. Clinical Features

Our sample average age of PD at diagnosis as well as the observed male predominance is consistent with prior literature. A history of non-potable water consumption along with exposure to pesticides and herbicides was reported in up to 40% of the patients. Previous exposure to pesticides and herbicides is associated with the development of PD [32]; similarly, exposure to welding and heavy metals such as manganese, copper, iron and mercury have been suggested to increase risk of PD. In this study, 22.1% and 11.6% of the patients reported frequent exposure to welding and other heavy metals, respectively.

The majority of our patients fulfilled Group A Gelb criteria while up to 60% also reported at least one of Group B symptoms, being the most frequent dystonia, falls and dysphagia. Common non-motor manifestations such as hiposmia, sleep disorders and depressive/anxious mood were seen in more than half our PD cases.

Performing regular physical activity correlated with lower ON UPDRS scores in spite of increasing age. Physical activity has been established as a possible protective factor for incident Parkinsonism [33], our data would be suggesting that physical activity could determine reduced severity of disease, specifically concerning motor features. Although exercise has not proven to slow the progression of akinesia, rigidity and gait disturbances, it promotes a feeling of physical and mental wellbeing and at the same time it can alleviate rigidity related pain and improve patients motor [34] and nonmotor symptoms [35].

Increasing age, coffee consumption, hallucinations, falls and mood disorders along with reduced years of education significantly correlated with worse MoCA scores. Older age and duration of PD are determinant risk factors for incidence of dementia in PD [36]. Furthermore, hallucinations have been established as risk factors for cognitive impairment [36, 37] along with gait disturbances (manifested by falls) [38] and depression [39]. Reduced education years also have been proposed as a risk factor for cognitive impairment in patients with PD [40]. Poor global cognition has been previously associated with a higher hazard of incident parkinsonism [41]. There have been inconsistent findings regarding the effects of coffee consumption on specific cognitive domains. It has been suggested to be in association with improved executive performance but smaller hippocampal volume and worse memory function [42]; however, this association is not sustained when cognition is analyzed longitudinally. Other literature suggested that coffee consumption might be slightly beneficial on memory without a dose response relationship [43]. Recent large-scale genetic analysis using mendelian randomization did not find any evidence supporting any beneficial or adverse long-term effect of coffee consumption on global cognition or memory function [44] or AD incidence [45]. To our knowledge, there is no literature evaluating the effect on cognition of coffee consumption, specifically for PD patients. Our findings suggest a possible deleterious effect that should be further explored in this population.

### 4.2. Genetic assessment

After sequence coding familial PD and atypical parkinsonism-associated genes including *GBA, SNCA, VPS35, LRRK2, GCH1, PRKN, PINK1, DJ-1, VPS13C, ATP13A2* and correcting for multiple comparisons, burden and SKAT-O analyses did not show an association of any of the tested genes and PD. We also did not identify any homozygous or compound heterozygous pathogenic variants in any of these genes. Rare variants in *LRRK2* were nominally associated with PD, observing only in affected individuals, six of these rare nonsynonymous variants, all located between amino acids p.1620-1623 in the COR domain of *LRRK2. LRRK2* encodes a multiple domain protein that includes a Roc-COR tandem domain, a tyrosine kinase-like protein kinase domain and at least four repeat domains located within the N-terminal and C-terminal regions. The Roc-COR domain classifies the LRRK2 protein as part of the ROCO superfamily of Ras-like G proteins [46]. Mutations in *LRRK2* are the most common cause of late onset AD hereditary PD. Most frequently reported disease-causing mutations locate in the kinase domain (i.e. G2019S) increasing kinase activity; and in the Roc-COR tandem domain (i.e. R1441C/G and Y1699C) impairing its GTPase function. Alterations of both kinase and GTPase activity may mediate neurodegeneration in these forms of PD [47]. Of the six patients found to have nonsynonymous variants in the COR domain, two had first degree relatives with dementia, one had a second degree relative with PD and one had two sisters with PD diagnosed at a very young age (20 and 30 years old) (see Table 4).

Nonsynonymous *GBA* variants were identified in three individuals including one patient and two unaffected controls. While we cannot rule out that these healthy individuals will develop PD in the future, it is unlikely that *GBA* variants have a major role in PD among Costa Rican patients.

Finally, one PD patient carried a pathogenic variant, p.K224R in *GCH1* gene. *GCH1* encodes for GTP cyclohydrolase 1 which is a key enzyme for dopamine production in nigrostriatal neurons. Loss of function mutations such as p.K224R have been shown to cause Dopa-responsive dystonia (DRD), however variants in this gene, have also been implicated in PD, perhaps through regulation of *GCH1* expression [48, 49]. It has been suggested that late-onset DRD might present clinically with parkinsonism; or alternatively pathogenic *GCH1* mutations may predispose to both diseases and carriers will develop any of both depending on other genetic or environmental factors [50]. Our patient did not present clinical features suggestive of DRD and did not have any family history of PD.

Genome analysis from *Mestizo* populations in Latin America showed in Costa Rica a European, Native American and African admixture of 66.7%, 28.7% and 4.6%, respectively [51]. Therefore, we would have expected to observe a higher prevalence of mutations, similar to other European series reported. However, our sample is more representative of the metropolitan area where most of the patients were recruited, thus warranting in the future, for a more comprehensive study involving a wider and more representative population of the whole country. Particularly, including more patients from the non-metropolitan and coastal zones.

## 5. Conclusions

This is the first study that reports on sociodemographic, risk factors, clinical presentation and genetics of Costa Rican patients with PD. We observed a high prevalence of exposure to both risk factors (pesticides, herbicides, non-potable water, low education) as well as protective factors (tobacco and coffee intake). Regular physical activity significantly correlated with better UPDRS scores despite years of evolution of the disease. Increased years of education were significantly associated with better MoCA test scores, whereas the presence of hallucinations, falls and mood disorders correlated with a worse performance in MoCA test. Interestingly, coffee consumption also correlated significantly with worse MoCA test scoring.

We did not find an association of any of the tested familial PD and atypical parkinsonism-associated genes including *GBA, SNCA, VPS35, LRRK2, GCH1, PRKN, PINK1, DJ-1, VPS13C, ATP13A2* and PD. We also did not identify any homozygous or compound heterozygous pathogenic variants in any of these genes. Rare variants in *LRRK2* were nominally associated with PD, six of these rare nonsynonymous variants, all located in the COR domain of *LRRK2*. One PD patient carried a pathogenic *GCH1* variant, p.K224R, further emphasizing the role of this gene in PD.

## Data Availability

Data is available upon reasonable request.

## Acknowledgements

We thank the participants for their contribution to the study. ZGO is supported by the Fonds de recherche du Québec–Santé Chercheur-Boursier award and is a Parkinson Canada New Investigator awardee. We thank Jennifer Ruskey, Sandra Laurent, Lynne Krohn, Uladzislau Rudakou, D. Rochefort, H. Catoire, V. Zaharieva and Dr. Ellen Sylvie-Mas for their assistance.

## Authors’ Roles

GTA and EY conceptualized the report and made substantial contributions to the design, drafting and revision of the work. EY and ZGO performed the genetic analysis. TLP, JRM, AGP, ZGO, KCC and JFT significantly contributed to drafting and critically reviewing the paper.

---

**Supplementary Table 1.** Average coverage of the 10 genes analyzed in this study along with the percentage of nucleotides covered at >15X and >30X for each gene.

| **Gene** | **Average coverage** | **Percentage of nucleotide passing >15x** | **Percentage of nucleotide passing >30x** |
| --- | --- | --- | --- |
| *DJ-1* | 829 | 100 | 100 |
| *ATP13A2* | 178 | 96 | 96 |
| *PINK1* | 671 | 97 | 97 |
| *GBA* | 758 | 100 | 97 |
| *SNCA* | 542 | 98 | 98 |
| *PARK2* | 799 | 98 | 98 |
| *LRRK2* | 470 | 97 | 97 |
| *GCH1* | 554 | 100 | 100 |
| *VPS13C* | 581 | 98 | 96 |
| *VPS35* | 502 | 96 | 96 |

**Supplementary Figure 1.**
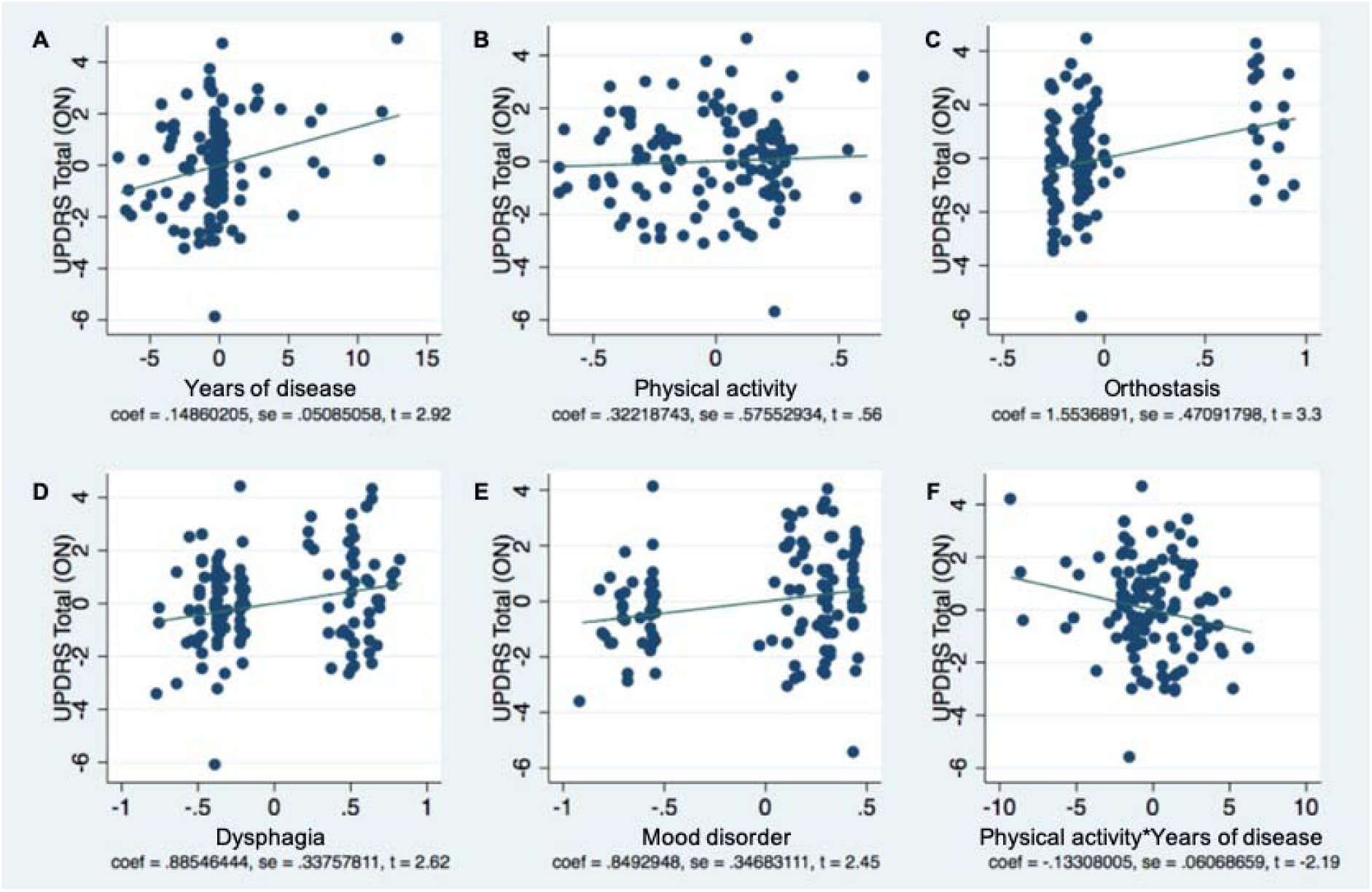
Multivariate linear regression model coefficients scatter plot assessing the relationship between clinical features of PD and total ON UPDRS score. In this model, increasing years of evolution of the disease (A) along with the presence of orthostasis (C), dysphagia (D) and mood disorders (E) significantly correlated with increased scores in total ON UPDRS. Furthermore, we found an interaction between performing regular physical activity and years of disease (F), where despite having increased years of disease, patients that performed regular physical activity still scored less in the total ON UPDRS.

**Supplementary Figure 2.**
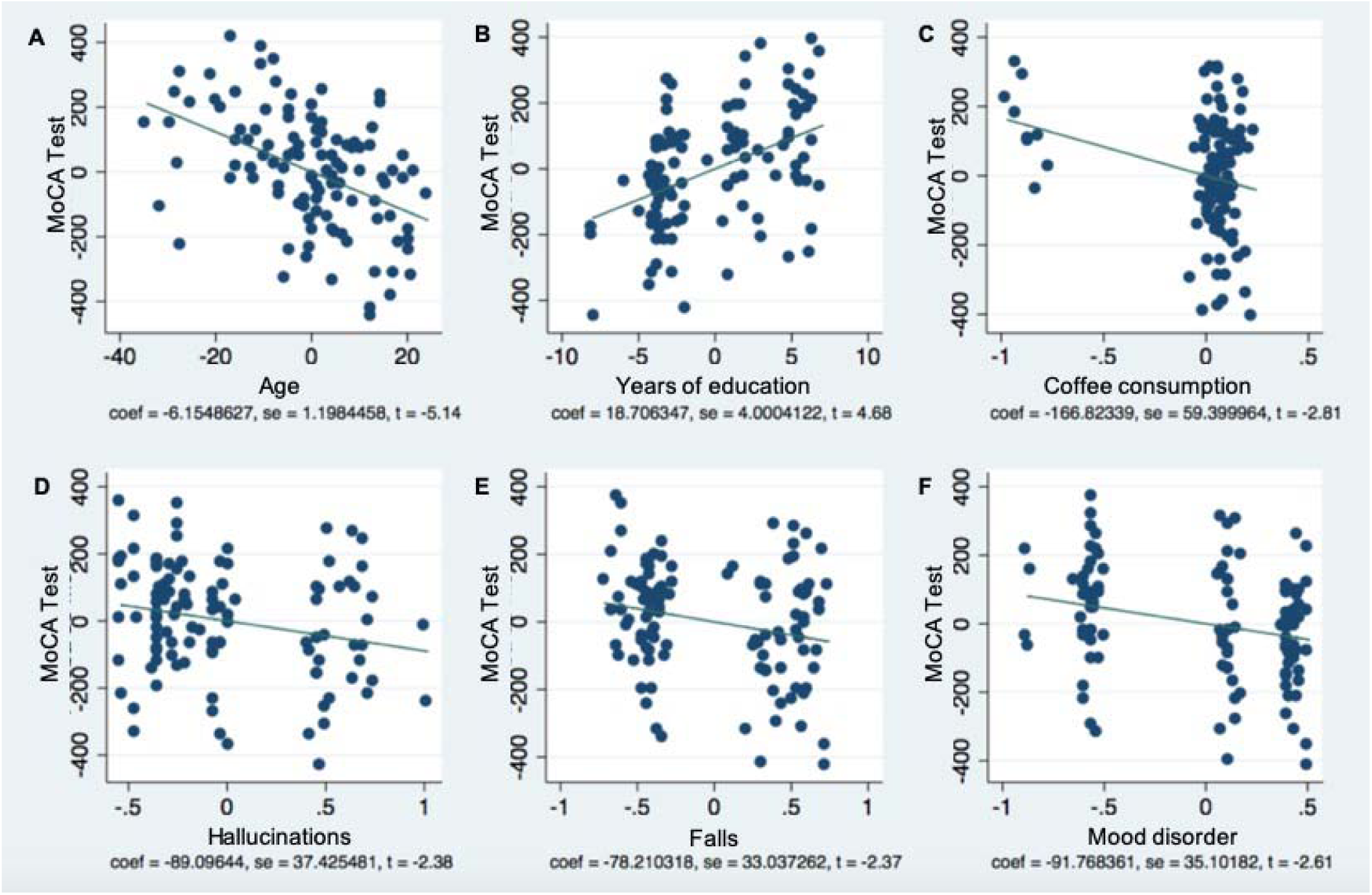
Multivariate linear regression model coefficients scatter plot for MoCA test scores. In this model, lower scores in MoCA test significantly correlated with increasing age (A), coffee consumption (C) and the presence of hallucinations (D), falls (E) and mood disorders (F), whereas increasing years of education (B) correlated with better MoCA scores.

